# Clinical evaluation of self-collected saliva by RT-qPCR, direct RT-qPCR, RT-LAMP, and a rapid antigen test to diagnose COVID-19

**DOI:** 10.1101/2020.06.06.20124123

**Authors:** Mayu Ikeda, Kazuo Imai, Sakiko Tabata, Kazuyasu Miyoshi, Nami Murahara, Tsukasa Mizuno, Midori Horiuchi, Kento Kato, Yoshitaka Imoto, Maki Iwata, Satoshi Mimura, Toshimitsu Ito, Kaku Tamura, Yasuyuki Kato

## Abstract

**Background:** The clinical performance of six molecular diagnostic tests and a rapid antigen test for severe acute respiratory syndrome coronavirus 2 (SARS-CoV-2) were clinically evaluated for the diagnosis of coronavirus disease 2019 (COVID-19) in self-collected saliva.

**Methods:** Saliva samples from 103 patients with laboratory-confirmed COVID-19 (15 asymptomatic and 88 symptomatic) were collected on the day of hospital admission. SARS-CoV-2 RNA in saliva was detected using a quantitative reverse-transcription polymerase chain reaction (RT-qPCR) laboratory-developed test (LDT), a cobas SARS-CoV-2 high-throughput system, three direct RT-qPCR kits, and reverse-transcription loop mediated isothermal amplification (RT-LAMP). Viral antigen was detected by a rapid antigen immunochromatographic assay.

**Results:** Of the 103 samples, viral RNA was detected in 50.5–81.6% of the specimens by molecular diagnostic tests and an antigen was detected in 11.7% of the specimens by the rapid antigen test. Viral RNA was detected at a significantly higher percentage (65.6–93.4%) in specimens collected within 9 d of symptom onset compared to that of specimens collected after at least 10 d of symptom onset (22.2–66.7%) and that of asymptomatic patients (40.0–66.7%). Viral RNA was more frequently detected in saliva from males than females.

**Conclusions:** Self-collected saliva is an alternative specimen diagnosing COVID-19. LDT RT-qPCR, cobas SARS-CoV-2 high-throughput system, direct RT-qPCR except for one commercial kit, and RT-LAMP showed sufficient sensitivity in clinical use to be selectively used according to clinical settings and facilities. The rapid antigen test alone is not recommended for initial COVID-19 diagnosis because of its low sensitivity.

**Key points:** Six molecular diagnostic tests showed equivalent and sufficient sensitivity in clinical use in diagnosing COVID-19 in self-collected saliva samples. However, a rapid SARS-CoV-2 antigen test alone is not recommended for use without further study.

## Introduction

Coronavirus disease 2019 (COVID-19), which is caused by severe acute respiratory syndrome coronavirus 2 (SARS-CoV-2), was first reported in 2019, in Wuhan, China, and the World Health Organization subsequently declared it a pandemic [1, 2]. The large number of patients with COVID-19 during outbreaks is overwhelming the capacity of national health care systems; therefore, the quick and accurate identification of patients requiring supportive therapies and isolation is important for the management of COVID-19.

The quantitative reverse-transcription polymerase chain reaction (RT-qPCR) assay for SARS-CoV-2 using upper and lower respiratory tract specimens (nasopharyngeal swab, throat swab, and sputum) is the gold standard for diagnosing COVID-19 [3]. Laboratory developed tests (LDT) including RT-qPCR, a high-throughput RT-qPCR system (fully automated from RNA extraction to reporting of results without the need for highly skilled laboratory technicians), and direct rapid RNA extraction-free RT-qPCR kits (using a modified RT-qPCR master mix), have been widely used worldwide [4]. Other molecular diagnostic methods such as reverse-transcription loop mediated isothermal amplification (RT-LAMP) have also been reported as useful for diagnosing COVID-19 in point-of care testing [5, 6]. Recently, a SARS-CoV-2 rapid antigen test (RAT) (ESPLINE® SARS-CoV-2; Fuji Rebio Inc., Tokyo, Japan), which combines immunochromatography with an enzyme immunoassay to detect the viral nucleocapsid protein, has been approved by the Japanese government [7]. The RAT is beginning to be used for diagnosing COVID-19 in clinical settings because it does not require special equipment, it does not have a time-consuming protocol, and highly skilled laboratory technicians are not essential. Although these diagnostic tests are useful in the identification of patients with COVID-19, the process of collecting upper and lower respiratory tract specimens increases the risk of exposure to viral droplets and there is a patient burden [8]. Therefore, an alternative specimen, which can be self-collected, to diagnose COVID-19 is desirable for the clinical management of COVID-19 during this pandemic era [8].

The sensitivity of RT-qPCR on upper respiratory specimens has been reported lower (32% for pharyngeal swab, and 63% for nasopharyngeal swabs) than lower respiratory specimens (72% for sputum, and 93% for bronchoalveolar lavage fluid) [9]. Recently, several reports highlighted the clinical usefulness of RT-qPCR analysis of saliva specimens [10-15]. Saliva specimens can be easily collected by the patients themselves by spitting into a collection tube; thus, using saliva specimens can reduce the burden on a patient, reduce the risk of exposure to viral droplets for medical workers, and reduce the time and cost of the testing procedure [16]. However, the clinical usefulness of saliva specimens for diagnosing COVID-19 remains controversial because the diagnostic sensitivity also vary widely between 69.2 and 100% for COVID-19, and it has yet to be thoroughly evaluated due to the small sample size and lack of detailed clinical information [10-15, 17].

Here, we describe the clinical performance of various molecular diagnostic methods including LDT RT-qPCR, cobas SARS-CoV-2 high-throughput system, 3 direct RT-qPCR kits and RT-LAMP, and a commercial SARS-CoV-2 RAT on self-collected saliva specimens in diagnosing COVID-19.

## Materials and methods

### Patients and sample collection

Patients with COVID-19 were enrolled in this study after being referred to the Self-Defense Forces Central Hospital in Japan for isolation and treatment under the Infectious Disease Control Law in effect from Feb 11 to May 13, 2020. All patients were examined for the SARS-CoV-2 virus by RT-qPCR using pharyngeal and nasopharyngeal swabs collected at public health institutes or hospitals in accordance with the nationally recommended method in Japan [18]. On the day of admission, saliva specimens (∼500 μL) were self-collected by all patients by spitting into a sterile tube. All samples were stored at −80 °C until sample preparation. All sample preparation and sample analysis were conducted by SRL, Inc. (Tokyo, Japan).

### Sample preparation

Saliva specimens were diluted with phosphate-buffered saline at a volume 1–5 times in accordance with the consistency and mixed with a vortex mixer. The suspension was centrifuged at 20,000 × g for 30 min at 4 °C and the supernatant was used in the following molecular diagnostic and RAT.

### Detection of viral RNA by LDT RT-qPCR using the standard protocol

LDT RT-qPCR was performed according to the National Institution Infections Diseases (NIID) protocol which is nationally recommended for SARS-CoV-2 detection in Japan [18]. Viral RNA was extracted from 140 μL saliva specimens using a QIAsymphony™ RNA Kit (QIAGEN, Hilden, Germany) following the manufacturer’s instructions. RT-qPCR amplification of the SARS-CoV-2 nucleocapsid (N) protein gene was performed using the QuantiTect®Probe RT-PCR Kit (QIAGEN) with the following sets of primers and probe. N-1 set: forward primer 5’ –CAC ATT GGC ACC CGC AAT C - 3’, reverse primer 5’ – GAG GAA CGA GAA GAG GCT TG - 3’, probe 5’ - FAM – ACT TCC TCA AGG AAC AAC ATT GCC A - TAMRA- 3’ [19]. N-2 set: forward primer 5’ - AAA TTT TGG GGA CCA GGA AC - 3’, reverse primer 5’ - TGG CAG CTG TGT AGG TCA AC - 3’, probe 5’ - FAM - ATG TCG CGC ATT GGC ATG GA - TAMRA-3’ [18]. A positive result with either or both of the primer and probe sets indicated the presence of viral RNA.

### Detection of viral RNA by direct RT-qPCR method without RNA extraction

Direct RT-qPCR methods without RNA extraction were performed using three commercial kits: Method A, SARS-CoV-2 Direct Detection RT-qPCR Kit (Takara Bio Inc. Kusatsu, Japan); Method B, Ampdirect™ 2019 Novel Coronavirus Detection Kit (Shimadzu Corporation, Kyoto, Japan); and Method C, SARS-CoV-2 Detection Kit (Toyobo, Osaka, Japan) according to the manufacturers’ instructions. The kits B and C were used with the same primer sets as used in the LDT RT-qPCR method. The kit A was used with the primer sets recommended by Centers for Disease Control and Prevention (CDC) [20]. Processed saliva specimens were directly added to the RT-qPCR master mix and then to thermal cycling, directly. Methods A and C are quantitative, whereas method B is qualitative.

### Detection of viral RNA by automated RT-qPCR device

The cobas SARS-CoV-2 test (Roche, Basel, Switzerland) [7, 21] was performed on the RT-qPCR automated cobas 8800 system (Roche) [4]. Specimens (600 μL) were loaded onto the cobas 8800 with cobas SARS-CoV-2 master mix containing an internal RNA control, primers, and probes targeting the specific SARS-CoV-2 open reading frame (ORF) 1 gene (target 1) and envelope (E) gene (target 2). A cobas 8800 positive result for the presence of SARS-CoV-2 RNA was defined as “detected” if targets 1 and 2 were detected or “presumptive positive” if target 1 was not detected but target 2 was detected.

### Detection of viral RNA by RT-LAMP

RT-LAMP detection of SARS-CoV-2 was performed using a Loopamp® 2019-SARS-CoV-2 Detection Reagent Kit (Eiken Chemical, Tokyo, Japan) according to the manufacturer’s instructions. The reaction was conducted at 62.5 °C for 35 min with the turbidity measuring real-time device LoopampEXIA® (Eiken Chemical).

### Detection of SARS-CoV-2 viral antigen by rapid antigen test

RAT was performed using ESPLINE® SARS-CoV-2 (Fuji Rebio Inc) according to the manufacturer’s instructions. In brief, the sample for analysis was obtained by dipping a swab which was provided by a RAT kit into the saliva specimen and then into the sample preparation mixture provided by the kit. The mixture (200 μL) was added to the sample port of the antigen assay. Subsequently, 2 drops of buffer were added and the results were interpreted after a 30 min incubation.

### Definitions

The saliva sample collection day was defined as day 1. Symptomatic cases were subdivided into two groups [22]. Severe symptomatic cases were defined as patients showing clinical symptoms of pneumonia (dyspnea, tachypnea, saturation of percutaneous oxygen [SpO2] < 93%, and the need for oxygen therapy). Other symptomatic cases were classified as mild cases.

### Ethical statement

Written informed consent was obtained from each enrolled patient at the Self-Defense Forces Central Hospital. This study was reviewed and approved by the Self-Defense Forces Central Hospital (approval number 02-024) and International University of Health and Welfare (20-Im-002-2).

### Statistical analysis

Continuous variables with a normal distribution were expressed as mean (± SD) and with a non-normal distribution as median (IQR), and compared using the Student’s t-test and Wilcoxon rank-sum test for parametric and nonparametric data, respectively. Categorical variables were expressed as number (%) and compared by χ2 test or Fisher’s exact test. The Kruskal-Wallis test was used for nonparametric analysis with over three independent samples. Linear regression analysis was used to assess the relationship between each molecular diagnostic method. A two-sided *p* value < 0.05 was considered statistically significant. All statistical analyses were calculated using R (v3.4.0; R Foundation for Statistical Computing, Vienna, Austria [http://www.R-project.org/]).

## Results

### Sensitivity of molecular diagnostic tests and antigen test

In this study, 7 diagnostic tests for COVID-19 were compared across 103 saliva specimens self-collected by 103 patients (Table 1). Singleton test was conducted for each method. Among the molecular diagnostic tests, LDT RT-qPCR showed the highest sensitivity on analyzing the 103 saliva samples. The direct RT-qPCR Method B exhibited higher sensitivity than either Method A or C. Only 12 patients tested positive using the RAT.

**Table 1.**
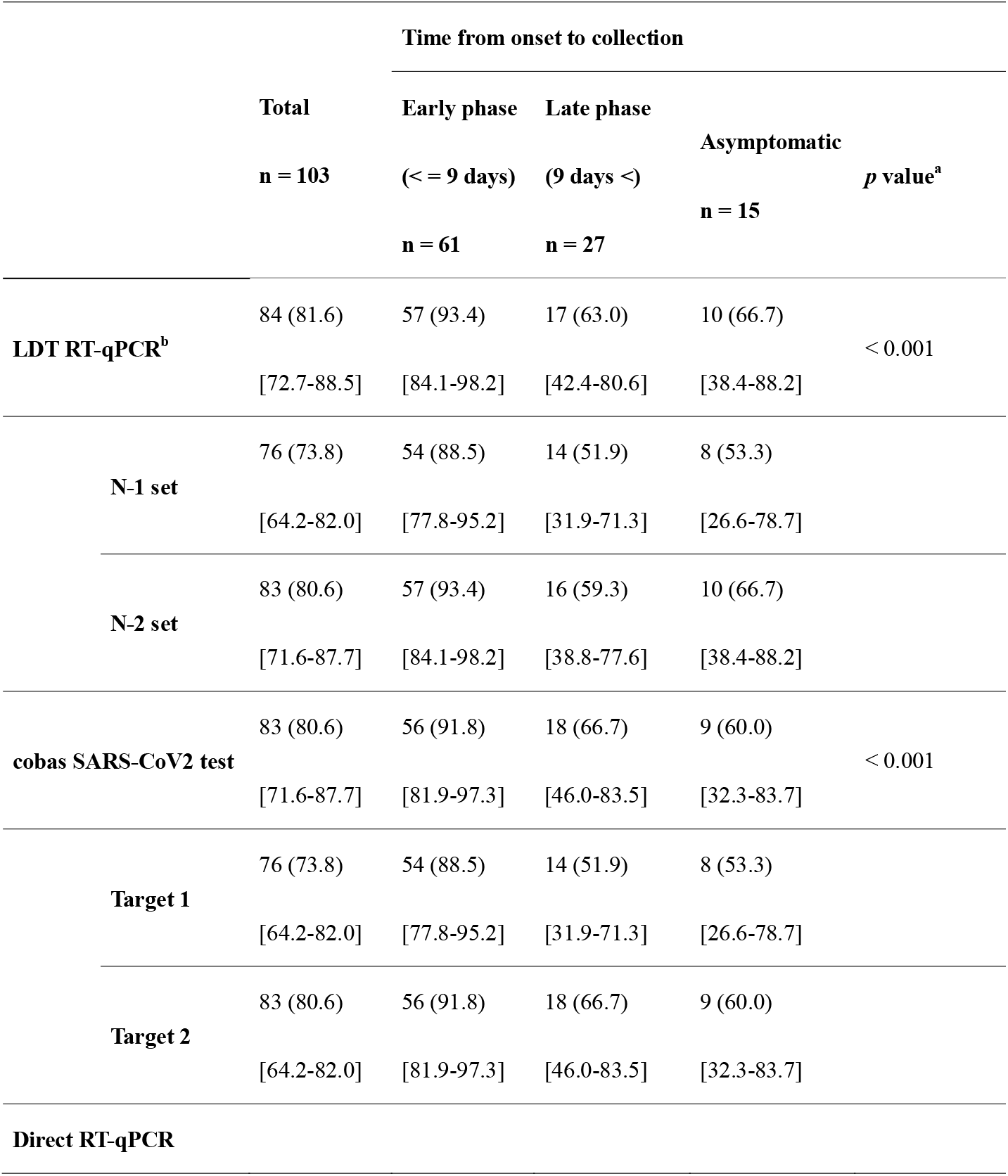
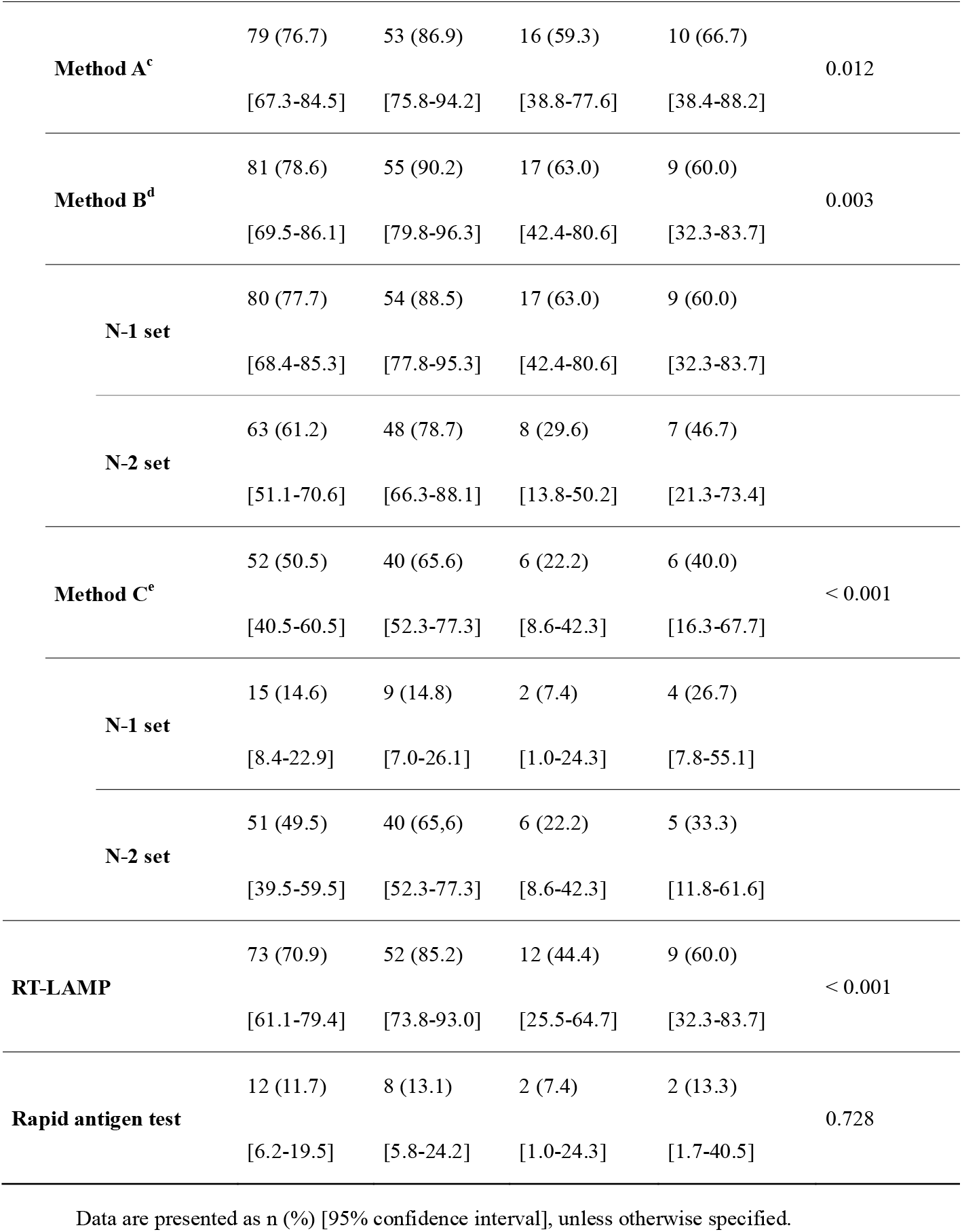
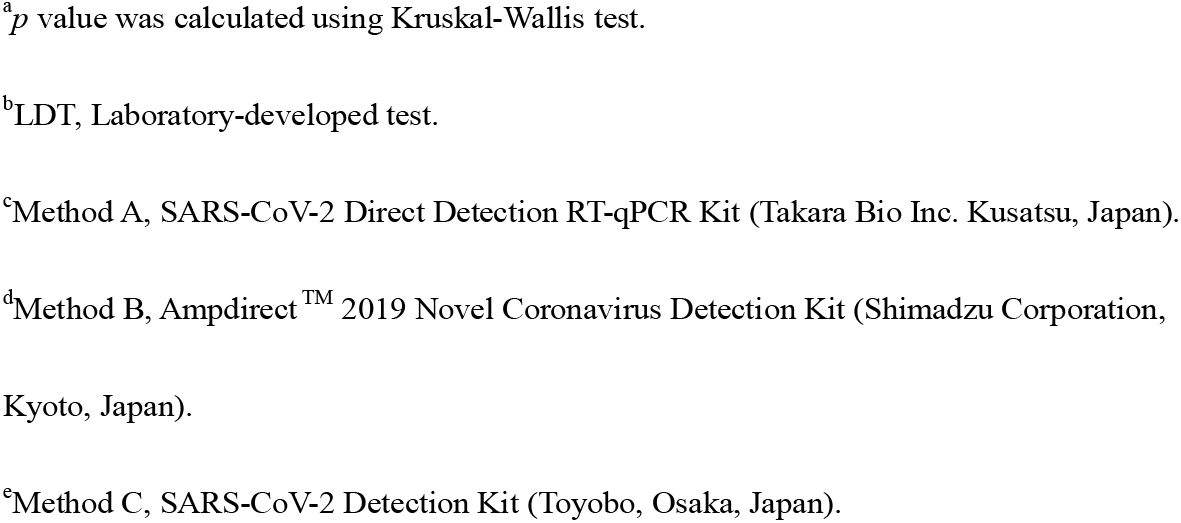
Summary of results of molecular diagnostic tests and rapid antigen test for COVID-19 on self-collected saliva samples.

|  | Total<br>n = 103 | Time from onset to collection |  | Asymptomatic<br>n = 15 | p value<sup>a</sup> |
| --- | --- | --- | --- | --- | --- |
|  |  | Early phase<br>(< = 9 days)<br>n = 61 | Late phase<br>(9 days <)<br>n = 27 |  |  |
| LDT RT-qPCR<sup>b</sup> | 84 (81.6) | 57 (93.4) | 17 (63.0) | 10 (66.7) | < 0.001 |
|  | [72.7-88.5] | [84.1-98.2] | [42.4-80.6] | [38.4-88.2] |  |
| N-1 set | 76 (73.8) | 54 (88.5) | 14 (51.9) | 8 (53.3) |  |
|  | [64.2-82.0] | [77.8-95.2] | [31.9-71.3] | [26.6-78.7] |  |
| N-2 set | 83 (80.6) | 57 (93.4) | 16 (59.3) | 10 (66.7) |  |
|  | [71.6-87.7] | [84.1-98.2] | [38.8-77.6] | [38.4-88.2] |  |
| cobas SARS-CoV2 test | 83 (80.6) | 56 (91.8) | 18 (66.7) | 9 (60.0) | < 0.001 |
|  | [71.6-87.7] | [81.9-97.3] | [46.0-83.5] | [32.3-83.7] |  |
| Target 1 | 76 (73.8) | 54 (88.5) | 14 (51.9) | 8 (53.3) |  |
|  | [64.2-82.0] | [77.8-95.2] | [31.9-71.3] | [26.6-78.7] |  |
| Target 2 | 83 (80.6) | 56 (91.8) | 18 (66.7) | 9 (60.0) |  |
|  | [64.2-82.0] | [81.9-97.3] | [46.0-83.5] | [32.3-83.7] |  |
| Direct RT-qPCR |  |  |  |  |  |
| **Method A<sup>c</sup>** | 79 (76.7) | 53 (86.9) | 16 (59.3) | 10 (66.7) | 0.012 |
|  | [67.3-84.5] | [75.8-94.2] | [38.8-77.6] | [38.4-88.2] |  |
| **Method B<sup>d</sup>** | 81 (78.6) | 55 (90.2) | 17 (63.0) | 9 (60.0) | 0.003 |
|  | [69.5-86.1] | [79.8-96.3] | [42.4-80.6] | [32.3-83.7] |  |
| **N-1 set** | 80 (77.7) | 54 (88.5) | 17 (63.0) | 9 (60.0) |  |
|  | [68.4-85.3] | [77.8-95.3] | [42.4-80.6] | [32.3-83.7] |  |
| **N-2 set** | 63 (61.2) | 48 (78.7) | 8 (29.6) | 7 (46.7) |  |
|  | [51.1-70.6] | [66.3-88.1] | [13.8-50.2] | [21.3-73.4] |  |
| **Method C<sup>e</sup>** | 52 (50.5) | 40 (65.6) | 6 (22.2) | 6 (40.0) | < 0.001 |
|  | [40.5-60.5] | [52.3-77.3] | [8.6-42.3] | [16.3-67.7] |  |
| **N-1 set** | 15 (14.6) | 9 (14.8) | 2 (7.4) | 4 (26.7) |  |
|  | [8.4-22.9] | [7.0-26.1] | [1.0-24.3] | [7.8-55.1] |  |
| **N-2 set** | 51 (49.5) | 40 (65.6) | 6 (22.2) | 5 (33.3) |  |
|  | [39.5-59.5] | [52.3-77.3] | [8.6-42.3] | [11.8-61.6] |  |
| **RT-LAMP** | 73 (70.9) | 52 (85.2) | 12 (44.4) | 9 (60.0) | < 0.001 |
|  | [61.1-79.4] | [73.8-93.0] | [25.5-64.7] | [32.3-83.7] |  |
| **Rapid antigen test** | 12 (11.7) | 8 (13.1) | 2 (7.4) | 2 (13.3) | 0.728 |
|  | [6.2-19.5] | [5.8-24.2] | [1.0-24.3] | [1.7-40.5] |  |

Data are presented as n (%) [95% confidence interval], unless otherwise specified.
<sup>a</sup>*p* value was calculated using Kruskal-Wallis test.
<sup>b</sup>LDT, Laboratory-developed test.
<sup>c</sup>Method A, SARS-CoV-2 Direct Detection RT-qPCR Kit (Takara Bio Inc. Kusatsu, Japan).
<sup>d</sup>Method B, Ampdirect<sup>TM</sup> 2019 Novel Coronavirus Detection Kit (Shimadzu Corporation,
Kyoto, Japan).
<sup>e</sup>Method C, SARS-CoV-2 Detection Kit (Toyobo, Osaka, Japan).

The Ct values for the N-1 and N-2 primer sets for the direct RT-qPCR Method C (35.5 ± 2.2 and 34.8 ± 2.4, respectively) were significantly (*p* < 0.001) greater than those for LDT RT-qPCR (32.8 ± 4.1 and 30.1 ± 4.4, respectively) (Figure 1). The mean detection time of RT-LAMP was 19.1 min (SD = 4.0) (Figure 1). A significant correlation was observed between RT-LAMP detection time and the Ct value of target 2 in the cobas SARS-CoV-2 test (*p* < 0.001) (Figure 2A). All patients with a positive SARS-CoV-2 RAT also tested positive for the six molecular diagnostic tests. The Ct value of target 2 in the cobas SARS-CoV-2 test was significantly lower in saliva samples that tested positive by RAT compared to that of samples that tested negative (25.4 ± 1.8 vs. 30.8 ± 2.7, respectively; *p* < 0.001; Figure 2B).

**Figure 1.**
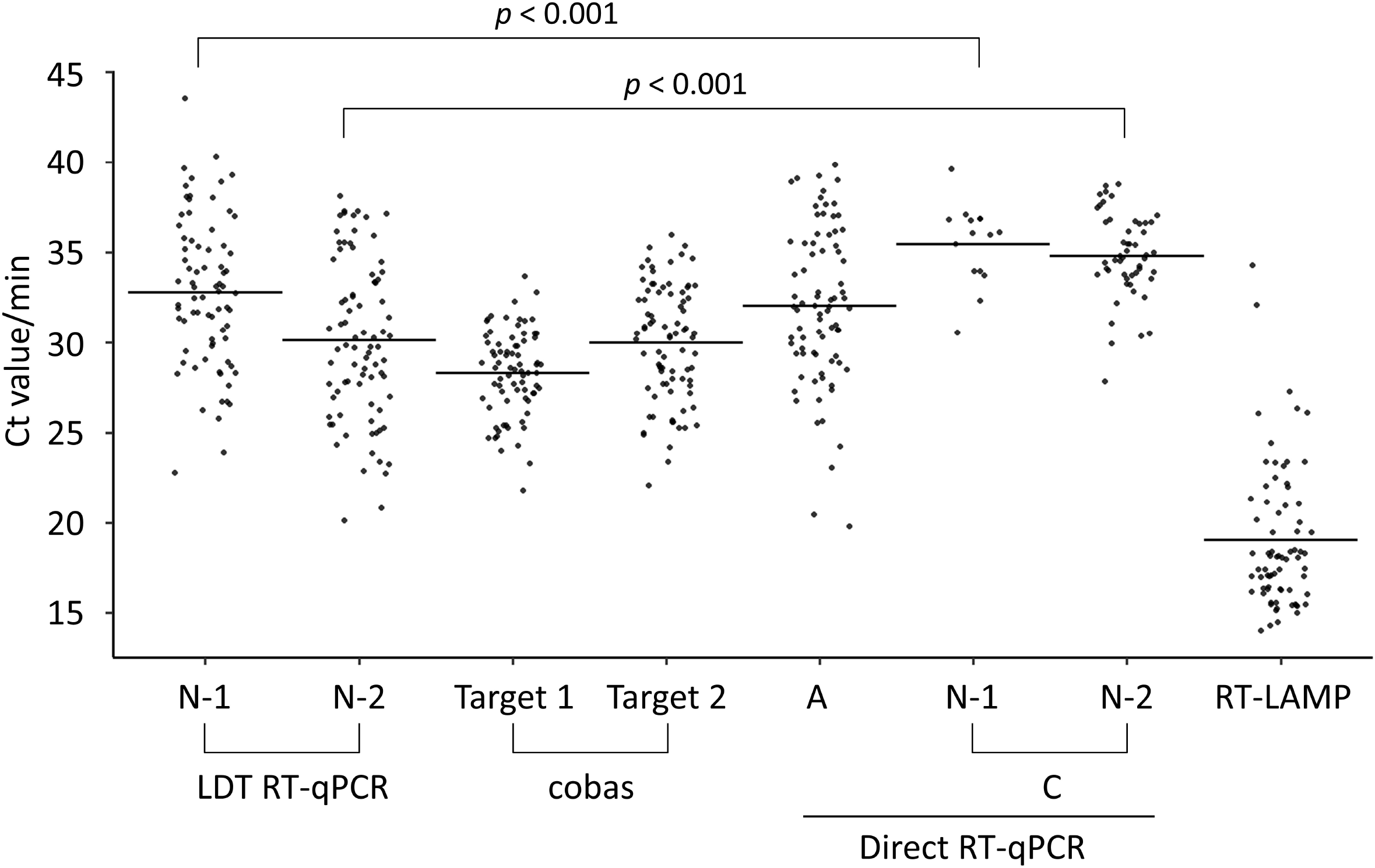
Ct value and detection time for each molecular diagnostic test of saliva specimens. Cycle threshold (Ct) value for each RT-qPCR primer set and detection time by reverse-transcription loop mediated isothermal amplification (RT-LAMP). Horizontal lines indicate the mean Ct value or detection time. *p* value was calculated using the Student’s t-test.

**Figure 2.**
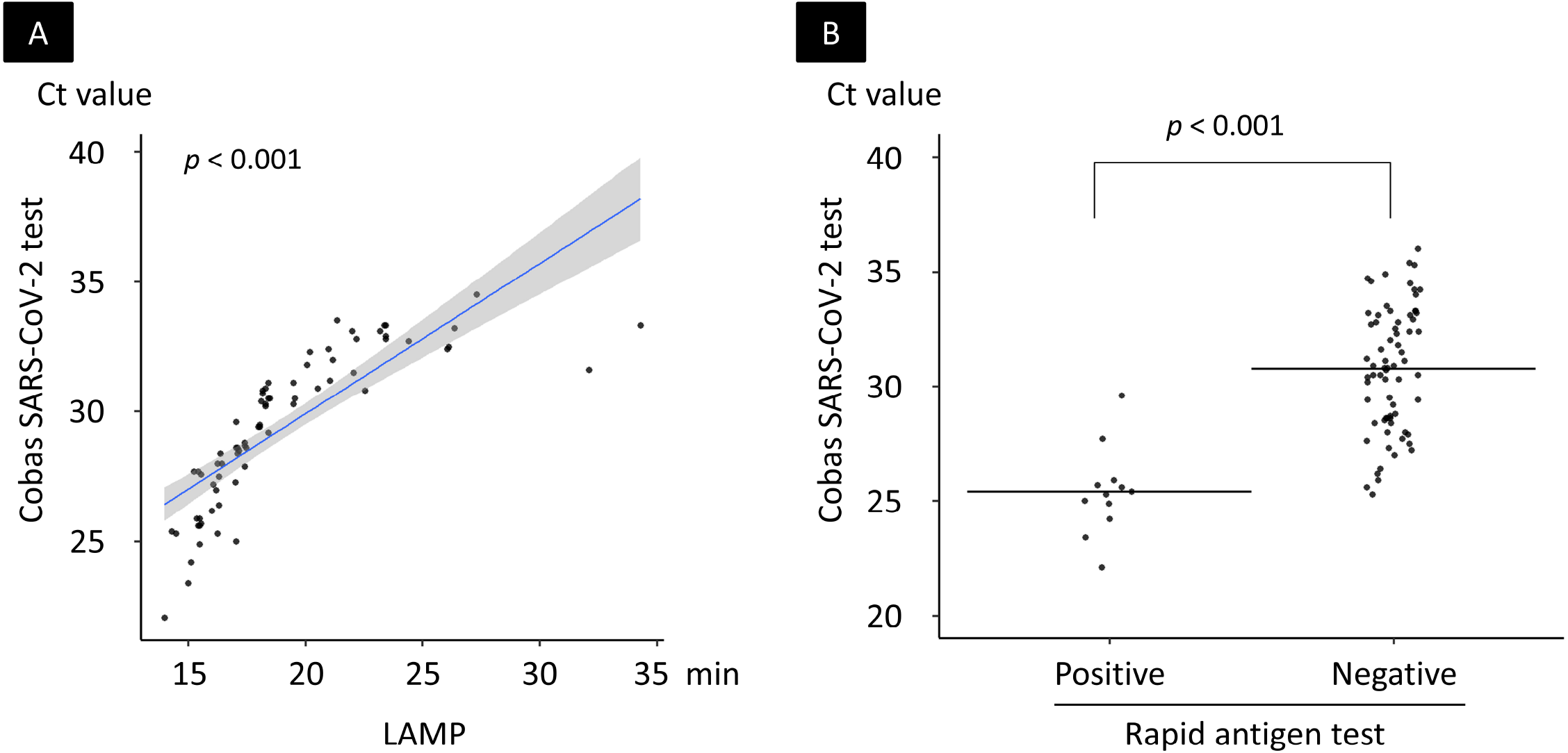
Relation of RT-qPCR, RT-LAMP, and rapid antigen test of saliva specimens. (A) Relation between detection time of reverse-transcription loop mediated isothermal amplification (RT-LAMP) and Ct value of target 2 (SARS-CoV-2 envelope gene) in cobas SARS-CoV-2 test. The blue slope line represents the fitted regression curve. The gray shadow indicates a 95% confidence interval around the regression curve. (B) Distribution of Ct value of target 2 in cobas SARS-CoV-2 test of saliva with positive and negative results. Horizontal lines indicate the mean Ct value. *p* value was calculated using the Student’s t-test.

### Effect of collection time on test sensitivity

On the day of admission, 15 patients (14.6%) who did not display any symptoms were classified as asymptomatic, whereas 88 patients (85.4%) were classified as COVID-19 symptomatic. Of the 88 symptomatic patients, saliva specimens were collected by 61 patients (69.3%) within 9 d from symptom onset (early phase of onset) and by 27 patients (30.7%) after 10 d from symptom onset (late phase of onset; Table 1). Samples from early phase, late phase, and asymptomatic patients tested positive by molecular diagnostic tests 65.6–93.4%, 22.3–66.7%, and 40.0–66.7%, respectively. The detection of saliva viral RNA was significantly higher in symptomatic patients who collected their saliva within 9 d from symptom onset than in saliva samples collected after 10 d from symptom onset and in saliva from asymptomatic patients (*p* < 0.01). There were no significant differences in prevalence of positive results by RAT among the three groups.

### Effect of clinical background on the prevalence of viral RNA in saliva

The baseline clinical characteristics of the 103 patients enrolled in this study are presented in Table 2. Briefly, patient age ranged from 18 to 87 years (median, 46 years; IQR, 38–63 years), and 66 (64.1%) patients were male. The time from symptom onset to sample collection was 1–14 d (median, 7 d; IQR, 6–10 d). The time from the initial RT-qPCR positive test to sample collection was 1–8 d (median, 4 d; IQR, 3–5 d). Of the 88 symptomatic patients, 72 (81.8%) and 16 (18.2%) were classified as mild and severe COVID-19, respectively.

**Table 2.** Effect of clinical background against the presence of viral RNA in saliva of 103 asymptomatic and symptomatic patients Presence of viral RNA in saliva.

|  | Total<br>n = 103 | Presence of viral RNA in saliva |  | *p* value<sup>a</sup> |
| --- | --- | --- | --- | --- |
|  |  | Positive<br>n = 84 | Negative<br>n = 19 |  |
| **Age (years)** | 48<br>[36–63] | 47<br>[39-67] | 44<br>[38-55] | 0.195 |
| **Sex** |  |  |  | 0.035 |
| **Male** | 66 (64.1) | 58 (69.0) | 8 (42.1) | - |
| **Female** | 37 (35.9) | 26 (31.0) | 11 (57.9) | - |
| **Disease activity** |  |  |  | 0.146 |
| **Asymptomatic** | 15 (14.6) | 10 (11.9) | 5 (26.3) | - |
| **Symptomatic** | 88 (85.4) | 74 (88.1) | 14 (73.7) | - |

Data are presented as n (%) or median [interquartile range], unless otherwise specified.
<sup>a</sup>*p* value was calculated using Wilcoxon rank-sum test for continuous variable, and $\chi^2$ test or
Fisher's exact test for categorical variables.

The effect of clinical background against the prevalence of viral RNA in saliva was analyzed using the results of LDT RT-qPCR, which had the highest sensitivity of all of the methods in this study. Among 103 patients, a significant male sex bias was noted in samples that tested positive for the virus compared to that of samples which tested negative (69.0% vs. 42.1%, respectively; *p* = 0.035) (Table 2). There were no significant differences in distribution by age or disease activity between patients detected or undetected with viral RNA (*p* > 0.05).

A summary of clinical symptoms and disease severity is shown for 88 symptomatic patients in Table 3. The disease symptom cough was observed in 41 of 74 patients (55.4%) with viral RNA in their saliva compared to 4 of 14 patients (28.6%) who did not test positive for viral RNA (*p* = 0.084). All severe patients (16/16, 100%) tested positive for viral RNA in their saliva, while 58 of 72 (78.4%) mild patients tested positive (*p* = 0.064).

**Table 3.** Effect of clinical background against the presence of viral RNA in saliva of 88 symptomatic patients.

|  | Symptomatic patients<br>n = 88 | Presence of viral RNA in saliva |  | *p* value<sup>a</sup> |
| --- | --- | --- | --- | --- |
|  |  | Positive<br>n = 74 | Negative<br>n = 14 |  |
| Age (years-old) | 46<br>[38-62] | 46<br>[38-60] | 44<br>[37-53] | 0.344 |
| Sex (male) |  |  |  | 0.011 |
| Male | 59 (67.0) | 54 (73.0) | 5 (35.7) | - |
| Female | 29 (33.0) | 20 (27.0) | 9 (64.3) | - |
| Disease severity |  |  |  | 0.064 |
| Mild | 72 (81.8) | 58 (78.4) | 14 (100) | - |
| Severe | 16 (18.2) | 16 (21.6) | - | - |
| Clinical symptoms |  |  |  |  |
| Fever | 73 (83.0) | 62 (83.8) | 11 (78.6) | 0.700 |
| Cough | 45 (51.1) | 41 (55.4) | 4 (28.6) | 0.084 |
| Malaise | 30 (34.1) | 25 (33.8) | 5 (35.7) | 1.000 |
| Headache | 25 (28.4) | 22 (29.7) | 3 (21.4) | 0.749 |
| **Diarrhea** | 20 (22.7) | 15 (20.3) | 5 (35.7) | 0.294 |
| **Sore throat** | 19 (21.6) | 15 (20.3) | 4 (28.6) | 0.491 |
| **Tachypnea** | 13 (14.8) | 13 (17.6) | - | 0.117 |
| **Dyspnea** | 8 (9.1) | 8 (10.8) | - | 0.346 |
| **Hypoxemia (SpO2 < 93 %)** | 10 (11.4) | 10 (13.5) | - | 0.353 |

Data are presented as n (%) or median [interquartile range], unless otherwise specified.
<sup>a</sup>*p* value was calculated using Wilcoxon rank-sum test for continuous variable, and $\chi^2$ test or
Fisher's exact test for categorical variables.

## Discussion

Here, we present evidence for the clinical usefulness of saliva specimens in diagnosing COVID-19. Previous studies reported that the sensitivity of RT-qPCR-analyzed saliva specimens initially collected from hospitalized patients was 69.2–100% for COVID-19 [10-15, 17]. The contradiction in sensitivity probably reflects differences in the clinical background and timing of sampling in each study. Becker et al, reported the lowest sensitivity (69.2%) using the clinical samples which were collected in late phase of onset [17]. On the other hands, Azzi et al, reported the highest sensitivity on saliva (100%) among hospitalized patients with severe and very severe disease [11]. In our study, the detection of viral RNA in saliva was significantly higher in samples collected in the early phase of symptom onset (within 9 d) compared to that in samples collected in the late phase of symptom onset (over 10 d), and in male patients versus female patients. Since the viral load of SARS-CoV-2 in saliva has been shown to decline from symptom onset [12], saliva specimens should be collected during the early phase of symptom onset to increase sensitivity.

SARS-CoV-2 uses the angiotensin-converting enzyme 2 (ACE2) on host cells found in the salivary gland and tongue tissues as well as nasal mucosa, nasopharynx, and lung tissue [23, 24] as a cell receptor to invade human cells [25]. Chen et al. collected saliva directly from the opening of the salivary gland and showed that SARS-CoV-2 can infect salivary glands [26]. The whole saliva flow rate is known to be higher in males than in females since it is associated with the size of the salivary gland [27]. The difference in saliva flow rate may affect the viral load in saliva and be associated with the difference in diagnostic sensitivity between males and females. We did not observe significant differences in disease severity or clinical symptoms between patients detected with or without saliva viral RNA; however, the prevalence of severe disease and the symptom of cough were frequently observed in patients detected with viral RNA in their saliva. Regarding disease activity, the presence of viral RNA was detected in more than 50% of the asymptomatic patients. These findings support previous studies reporting the presence of viral RNA in saliva of both symptomatic and asymptomatic patients [14]. Therefore, our findings revealed that saliva, collected in the early phase of symptom onset, is a reliable and practical source for the screening and diagnosing of COVID-19.

The clinical performance of direct RT-qPCR kits and RT-LAMP, and any correlation with RT-qPCR results were not well evaluated because of the small number of clinical specimens collected from patients in previous studies [5, 6, 28]. The sensitivity of RT-LAMP for SARS-CoV-2 using upper and lower respiratory tract specimens has been reported as equivalent to RT-qPCR [5, 6, 28]. However, our results indicate that the sensitivity of RT-LAMP is inferior to LDT RT-qPCR and cobas SARS-CoV-2 test for COVID-19 in saliva specimens. Direct RT-qPCR kits without an RNA extraction process can reduce the time, cost, and human resources needed to conduct the assay. However, we showed that there is a large difference in sensitivity among the direct RT-qPCR kits. It is necessary to pay attention to the false-negative results of RT-LAMP and direct RT-qPCR kits, especially when testing saliva samples. In clinical settings with limited medical and human resources, using RT-LAMP and direct RT-qPCR kits are options for screening and diagnosing COVID-19 because of their simplicity.

In comparison with molecular diagnostic tests, the SARS-CoV-2 RAT of saliva specimens showed low sensitivity. The sensitivity of RAT is still unclear, not only when using saliva samples but also when using nasopharyngeal swab specimens [7]. The experiment to compare the sensitivity of RT-qPCR and RAT prior to the approval as in vitro diagnostic test by Japanese government showed that sensitivity of RAT was 66.7% (16/24 patients) for nasopharyngeal swabs; furthermore, low sensitivity specimens contained a low viral copy number (50% sensitivity [6/12 patients] for specimens containing < 100 copies/test) [7]. Our findings also suggest that the RAT requires a high viral copy number to get positive result. This kit was not originally compatible with saliva specimens and the freeze-thaw process may have affected sensitivity. Improvements in sample preparation may increase its sensitivity.

Our study had several limitations. First, the saliva specimens were collected from patients 3 d (median) after receiving their first positive RT-qPCR result from analysis of upper respiratory specimens. Directly comparing the sensitivity between saliva and other upper or lower respiratory specimens is difficult in our study design because the viral load in the clinical specimens vary with time [13]. Second, although the high specificity of RT-qPCR for SARS-CoV-2 has been confirmed [6, 18, 28-31], the specificities should be analyzed by also using saliva from non-COVID-19 patients. Further studies are warranted to determine the usefulness of saliva specimens for screening and diagnosing COVID-19.

## Conclusions

Self-collected saliva in the early phase of symptom onset is an alternative specimen diagnosing COVID-19. LDT RT-qPCR, cobas SARS-CoV2 high-throughput system, direct RT-qPCR kits except for one commercial kit, and RT-LAMP showed equivalent and sufficient sensitivity in clinical use and can be selectively used according to the clinical setting and facilities. The rapid SARS-CoV-2 antigen test alone is not recommended for use at this time due to low sensitivity.

## Data Availability

The raw data used in this study is available from the corresponding author upon reasonable request.

## Conflict of interest

The authors declare that they have no conflicts of interests.

## Funding

This work was supported by the Health, Labour and Welfare Policy Research Grants, Research on Emerging and Re-emerging Infectious Diseases and Immunization [grant number 20HA2002].

## Acknowledgments

We thank clinical laboratory technicians at the Self-Defense Forces Central Hospital for sample collecting, and everyone involved in the COVID-19 Task Force at the Self-Defense Forces Central Hospital, and members who were assembled from other institutes of the Japan Self-Defense Forces.

## Authors’ contributions

YK and KI, study conception and design; MI and ST, collecting data and performing data analysis; MI, KI, KM, KT and YK manuscript drafting and editing; NM, TM, MH, KK, YI and MI, manuscript revision; SM, TI and KT, study supervision. All authors have read and approved the final manuscript.

## Notes

### Competing Interest Statement

The authors have declared no competing interest.

### Author Declarations

This study was reviewed and approved by the Self-Defense Forces Central Hospital (approval number 02-024) and International University of Health and Welfare (20-Im-002-2).

